# Chloroform associated with bone mineral density and bone mineral content in adults: a population-based cross-sectional research

**DOI:** 10.1101/2023.08.03.23293595

**Authors:** Lin Li, XK Liu, Xia Zhang, Yan Zhang, Qing Li, HF Geng, Li Shi, Ben Wang, QQ Qiu, TP Yu, YQ Sang, LY Wang, Wei Xu, Jun Liang

## Abstract

**Background:** According to a recent cross-sectional study, reduced Bone Mineral Density (BMD) in U.S. adults is associated with co-exposure to several Volatile Organic Compounds (VOCs), and contact with VOCs increases the risk of developing osteoporosis. However, the relationship between chloroform (an essential VOC component) and BMD remains unclear. Consequently, we aimed to explore the relationship between chloroform alone and Bone Mineral Density or Bone Mineral Content (BMD or BMC).

**Methods:** Herein, 2,553 individuals aged 18 and above from the National Health and Nutrition Examination Surveys (NHANES) in 2009-2010, 2013-2014, and 2017-2020, were included. We employed two independent t-tests and multi-linear regression models to statistically assess the relationship between chloroform exposure and BMD/BMC in the spine and femoral area.

**Results:** A “V”-shaped correlation between chloroform exposure and BMD/BMC was observed in the unadjusted model, particularly in the Ward’s triangle and femoral neck as a whole. A negative correlation was specifically observed for the Ward’s triangle BMD/BMC and L4 BMD/BMC. On the other hand, in the adjusted model, a dominantly negative correlation between the L4 BMC and chloroform exposure was observed over a range of exposure levels. The subgroup analysis revealed a negative correlation between chloroform concentrations and BMC in the femur and spine, especially in women and the 65-80 age population.

**Conclusion:** Our study revealed a “V” shaped correlation between chloroform and BMD/BMC of the femur and spine in U.S. adults. This finding highlights the fact that prolonged exposure to chloroform may increase the risk of developing osteoporosis.

## Introduction

Osteoporosis is a frequent bone disease characterized by reduced bone volume, microstructural degeneration, and fragility fractures [1]. Fragility fractures, particularly in the most vulnerable areas, including the vertebrae, hip, and forearm, are one of the severe complications of osteoporosis. According to reports [2,3], fragility fractures can cause emotional distress, reduced quality of life, and an increased socioeconomic burden. Approximately 79,000 people in the United Kingdom suffer hip fractures annually, with the cost in 2010 estimated at £3.5 billion and expected to rise to £5.5 billion each year by 2025, implying that osteoporosis will be a significant challenge in the future and its burden on the medical community must be removed [4]. According to osteoporosis research, enhancing Bone Mineral Density (BMD) is closely linked to decreasing the risk of vertebral and non-vertebral fractures [5]. Many factors, including aging, femininity, estrogen metabolism disturbances, and physical inactivity, are critical in lowering BMD and have been implicated in the pathogenesis of osteoporosis [6,7].

People are frequently exposed to Volatile Organic Compounds (VOCs) such as chloroform, bromoform, dibromochloromethane, and dichlorobromomethane. The increased contact is due to the widespread use of disinfectants, as these VOCs are produced when disinfectants react with organic substances in water sources [8, 9, 10].

Compared to normal mice, the U.S. National Toxicology Program noted an increased incidence of sternal dysplasia in bromoform-exposed mice [11]. Other studies discovered a connection between VOC exposure and bone-healing-related chemokines, with a downregulated osteoblast chemokine CXCL12 expression after exposure to benzene or its derivatives, and that CCL2 (a chemokine essential for osteoclast formation) production was induced by chlorobenzene exposure [12, 13, 14]. These findings imply that VOCs could impact BMD levels by affecting osteoblast activity. Additionally, individual and combined VOC exposure was found to be associated with lower BMD in U.S. adults, and Alkaline Phosphatase (ALP), Body Mass Index (BMI), Fasting Insulin (F.I.), and High-Density Lipoprotein (HDL) were shown to be intermediaries in VOC and BMD co-exposure [15]. Chloroform, a significant member of VOCs, is strongly absorbed through the pulmonary, oral, and dermal pathways and is linked with several acute and chronic illnesses, including neurological, respiratory, hepatic, and renal dysfunctions, reproductive abnormalities, and digestive or urinary malignancies [16, 17]. However, the connection between blood chloroform and BMD/BMC was rarely investigated alone in past studies.

This research aimed to explore the relationship between blood chloroform and BMD/BMC using data from the National Health and Nutrition Examination Survey (NHANES). We, therefore, obtained femur BMD/BMC and spine BMD/BMC datasets published in 2009-2010, 2013-2014, and 2017-2020 from NHANES.

## Materials and methods

### Study design and participants

Herein, data from the NHANES, a cross-sectional survey of more than 5,000 respondents conducted yearly by the Center for Disease Control and Prevention’s (CDC) Center for Health Statistics, was used. We specifically used NHANES data from 2009-2010, 2013-2014, and 2017-2020. We selected 22,333 adults (of whom 6,227 had blood chloroform data) from 36,272 available subjects. Subsequently, 2637 individuals with femur and spine Dual-Energy X-ray Absorptiometry (DEXA) data were selected. After excluding patients with a history of osteoporosis treatment, 2553 subjects aged ≥18 years were included in the final statistical analysis. **Fig1** depicts the flowchart for participant exclusion. (**Fig1:** Flowchart for Screening Research Subjects)

### Variables

In this study, we used blood chloroform concentration as the exposure variable. Chloroform concentration in the blood was analyzed using Capillary Gas Chromatography (G.C.) and Mass Spectrometry (M.S.) with Selected Ion Monitoring (SIM) detection and isotope dilution. The chloroform detection limits ranged from 0.00148 to 0.602 ng/ml, and for lower LLOD (Lower Limit of Detection) values, an interpolated LLOD value divided by the square root of 2 was used. The dependent variables were the femoral and spinal Bone Mineral Density (BMD) and Bone Mineral Content (BMC). In Shepherd’s laboratory, Dual-Energy X-ray Absorptiometry (DXA) was used to measure the spine and femur BMC, whereas the Hologic software APEX v4.0 (Hologic) sector beam densitometry was used to scan and calculate the spine and femur BMC and BMD. All variables included as covariates in the study are listed below:

Gender (male, female), Race/ethnicity (Mexican-American, Non-Hispanic white, Non-Hispanic black, other Hispanic, other race), Household Poverty-Income Ratio (PIR by poverty line: values 0 to 0.99 indicate below poverty, ≥1.00 indicate poverty or above poverty)[18], Excessive alcohol consumption (>1 drink /day for women or >2 drinks /day for men (in the past 12 months)), Hypertension (blood pressure ≥140/90 mmHg or taking anti-hypertensive medication), Diabetes mellitus (self-reported diabetes history, Fasting Blood Glucose (FPG) level ≥7.0 mmol/L, HbA1c ≥6.5% or taking anti-diabetic medication), Arthritis, Cardiovascular disease (self-reported congestive heart failure, coronary artery disease, heart attack or stroke history), Thyroid problems (self-reported hyperthyroidism, hypothyroidism, thyroiditis), COPD(Chronic Obstructive Pulmonary Disease), Smoking (self-described as having smoked ≥100 cigarettes in a lifetime), History of osteoporosis treatment and osteopenia (Bone Mineral Density (BMD) T-score ≤ -2.5 in lumbar spine or femoral neck) [19]. Age, body weight, abdominal circumference, and BMI were all continuous covariates in our analysis. The blood chloroform, BMD, BMC, and covariance data were obtained from the NHANES website.

### Statistical analysis

Statistical analyses were conducted using R software (version 3.6.1), IBM SPSS version 25(IBM, Armonk, New York, USA), and GraphPad Prism9. The mean and Standard Deviation (S.D.) values were used for variables with continuous features, whereas categorical variable represented frequency. The Spearman correlation test was used to assess differences between the classification variables in the exposure group. The Kruskal-Wallis test or one-way ANOVA was used to examine differences in continuous variables between groups. Blood chloroform levels were grouped into tertiles (tertile 1: <33%, tertile 2: 33-66%, tertile 3: >66%) and univariate and multivariate linear regression models were tested to analyze the relationship between the different blood chloroform categories. We adjusted the confounding variables in the multi-linear regression model to investigate the correlation between blood chloroform levels and BMD/BMC (femoral and spine). These included Model 1 (the unprecedented model), Model 2 (for gender, age, race, and BMI adjustment), and Model 3 (for gender, age, race, BMI, osteoporosis, COPD, and thyroid problems adjustments, and further adjustment for arthritis, smoking and excessive drinking). Regression analysis results were expressed as β (beta) coefficients, 95% confidence intervals, and statistically significant P values (P values<0.05).

## Results

### Subjects characteristics

This study had 2553 participants (1288 women and 1265 men). Based on the femoral and spinal DEXA data, the average age of the participants and the average chloroform levels in their whole blood was 48.83 years and 0.0171 ± 0.0182 ng/mL (ranging from 0.0021 to 0.1790 ng/mL), respectively. The weighted characteristics of these participants based on the third-class classification are shown in **TableS1**. According to the femoral and spine DEXA data, differences among groups in baseline variables such as age, race, height, BMI, waist circumference, hypertension, arthritis, thyroid illnesses, and smoking were statistically significant (P<0.05).

### Connection between chloroform and BMD

**Tables1-2** and **S2** illustrate the results. Blood chloroform concentration by tertile had a statistically significant relationship with total femoral BMD (P < 0.05); femoral neck BMD (P < 0.01); rotor BMD (P < 0.05); inter-rotor BMD (P < 0.05); Ward’s triangle BMD (P < 0.01); and L4 BMD (P < 0.01).

The multiple linear regression models and weighted univariate analyses are detailed in **Tables1-2**. All regressions were conducted before adjusting for some of the covariates (model 1), except for total spine BMD (β -0.0020; 95% CI -0.0051, 0.0011), L1 BMD (β 0.0004; 95% CI -0.0030, 0.0039), L2 BMD (β - 0.0024; 95% CI -0.0057, 0.0009), and L3 BMD (β -0.0016; 95% CI -0.0048, 0.0016). A statistically significant correlation existed between increased blood chloroform concentration and total femoral BMD, femoral neck BMD, rotor BMD, inter-rotor BMD, Ward’s triangle BMD, and L4 BMD (P<0.05). The L4 BMD was negatively correlated with blood chloroform levels after adjusting for some covariates (model 2). After adjusting for more relevant covariates simultaneously (model 3), L4 BMD decreased with rising blood chloroform levels, but not at other sites.

### Connection between chloroform and BMC

Detailed results are presented in **Tables 1-2** and **S2**. Weighted univariate and multiple linear regression analyses are illustrated in Tables 1-2. In model 1, blood chloroform levels were categorized by tertile, except for rotor BMC (β -0.0023; 95% CI -0.0066, 0.0021), and L1 BMC (β -0.0004; 95% CI -0.0058, 0.0049). There was a negative correlation between increased blood chloroform concentrations and BMC at several sites, including total femoral BMC, femoral neck BMC, Intertrochanter BMC, Ward’s triangle BMC, total spine BMC, L2 BMC, L3 BMC, and L4 BMC. Additionally, increased blood chloroform concentrations in the femur and spine were negatively correlated with BMC (P < 0.05). The L3 BMC and L4 BMC were negatively correlated with blood chloroform levels after adjusting for some covariates (model 2). Finally, in model 3, L3 BMC and L4 BMC decreased with increasing blood chloroform concentration.

**Table 1:**
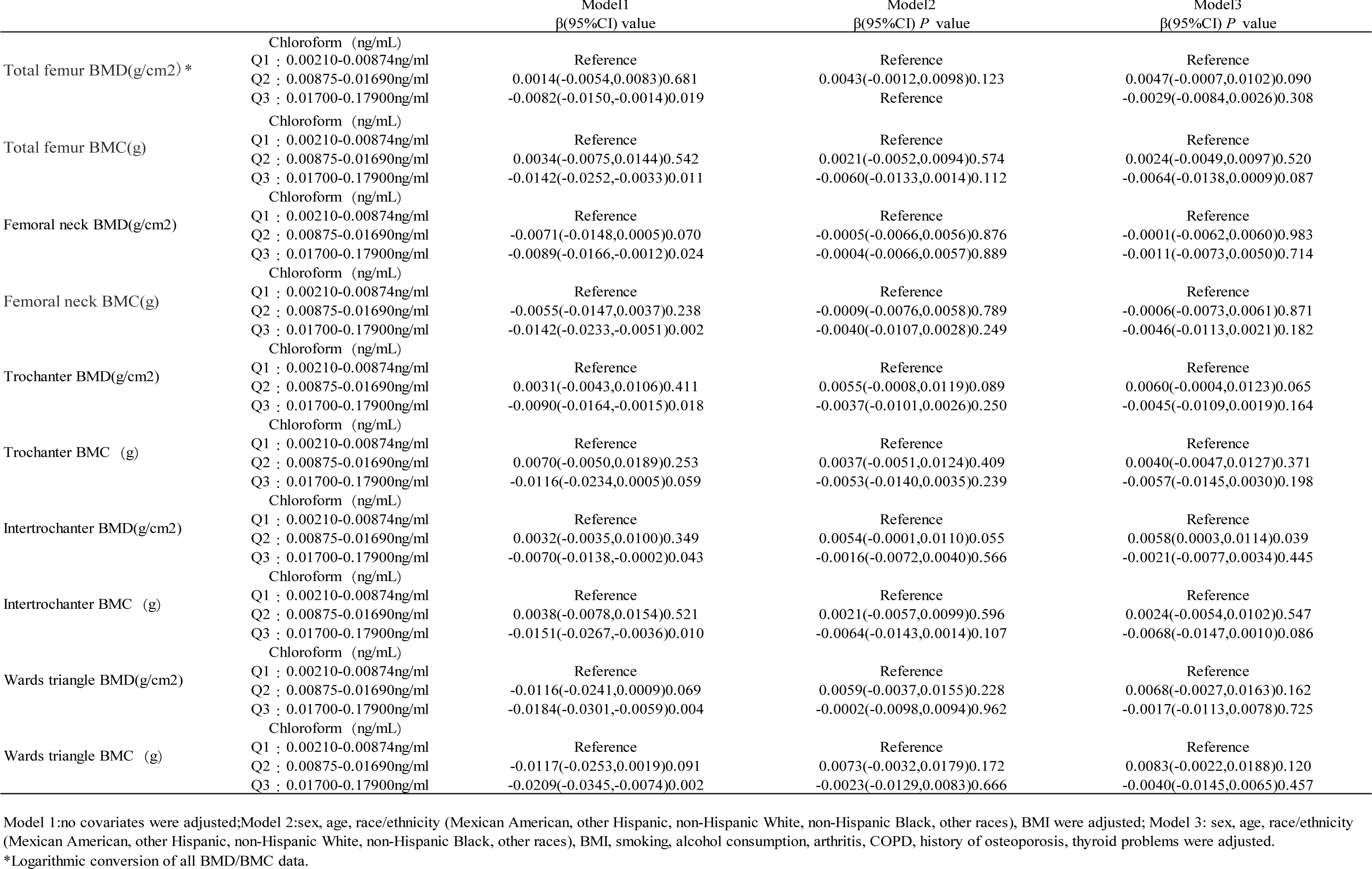
The relationship between blood chloroform and BMD/BMC in the femur.

**Table 2:**
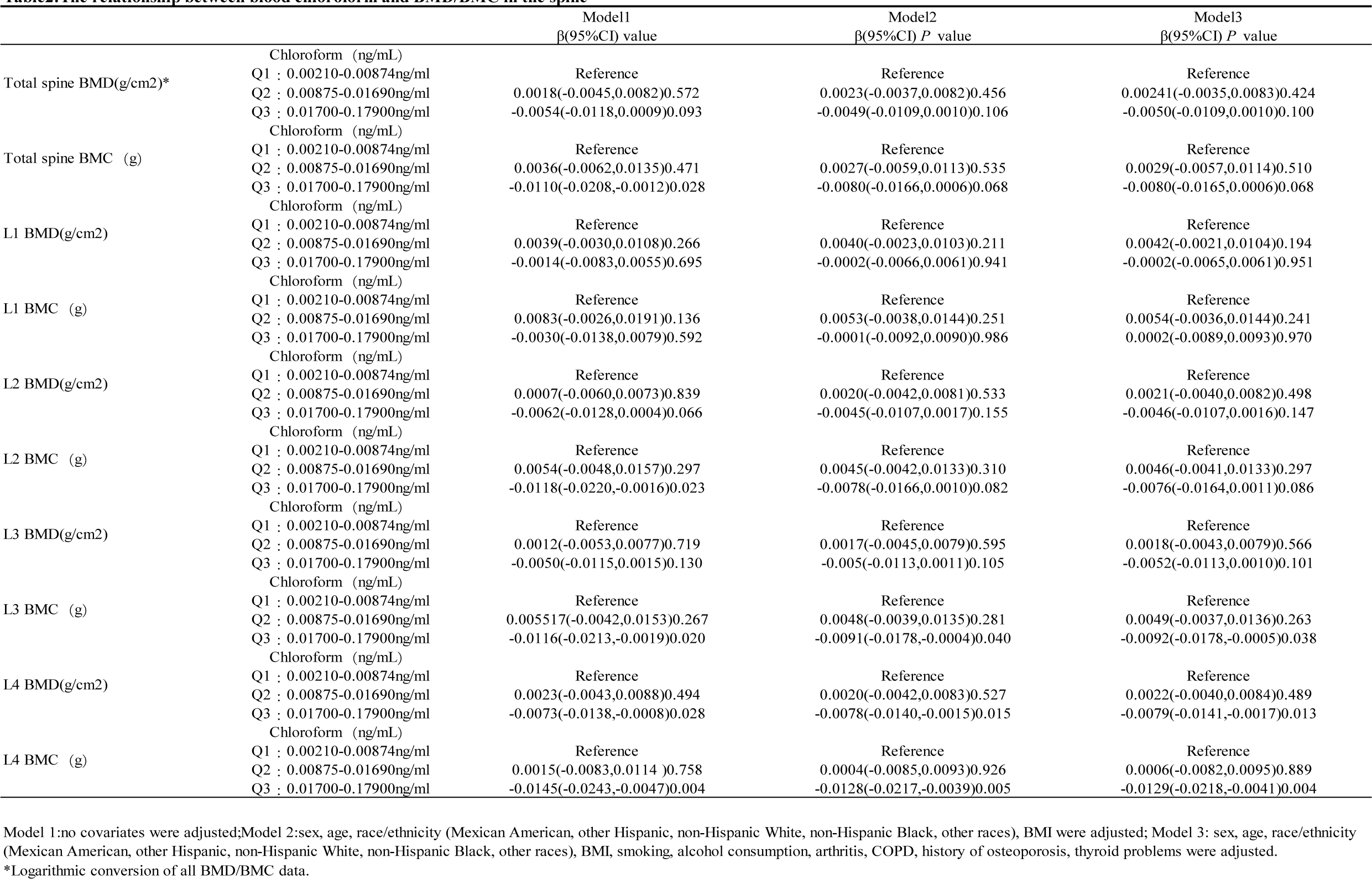
The relationship between blood chloroform and BMD/BMC in the spine.

### Subgroup analysis

The findings of subgroup analyses adjusted for associated covariates are shown in **Tables 3-5**. According to the Multiple regression analysis in the femoral area, the femoral neck BMC blood chloroform level decreased in the male gender (β -0.0059; 95 % CI -0.0115, -0.0002; P <0.05). According to the analysis by race as a subgroup, the femoral neck BMD/BMC in Non-Hispanic White and other Race-Including Multi-Racial subjects exhibited a negative correlation with blood chloroform concentration (P<0.05). In contrast, no meaningful correlation between blood chloroform and BMD or BMC was observed in Mexican-American, Non-Hispanic Black, and Other Hispanic subjects. A negative correlation between blood chloroform and femoral neck BMD/BMC was revealed by the results stratified by age but only in the 65-80 years-old participants (β -0.0100; 95% CI -0.0186, -0.0014; P<0.05); (β -0.0144; 95% CI -0.0234, -0.0054; P<0.05).

**Table 3.** Association of blood chloroform with femur neck BMD/BMC, stratified by sex, race/ethnicity and age.

| | femur neckBMD (g/cm<sup>2</sup>) | $\beta$(95%CI) value | femur neck (g) | $\beta$(95%CI) value |
| --- | --- | --- | --- | --- |
| Subgroup analysis stratified by gender |  |  |  |  |
| male | -0.0020(-0.0073,0.0033) | 0.454 | -0.0045(-0.0100,0.0011) | 0.114 |
| female | -0.0046(-0.0100,0.0008) | 0.094 | -0.0059(-0.0115,-0.0002) | 0.041 |
| Subgroup analysis stratified by race/ethnicity |  |  |  |  |
| Mexican American | -0.0020(-0.0098,0.0058) | 0.617 | -0.0041(-0.0139,0.0058) | 0.416 |
| Other Hispanic | -0.0052(-0.0163,0.0060) | 0.361 | -0.0079(-0.0215,0.0057) | 0.253 |
| Non-Hispanic White | -0.0075(-0.0135,-0.0015) | 0.015 | -0.0086(-0.0158,-0.0015) | 0.018 |
| Non-Hispanic Black | -0.0086(-0.0178,0.0006) | 0.0664 | -0.0104(-0.0211,0.0003) | 0.056 |
| Other Race—Including Multi-Racial | -0.0119(-0.0242,0.0003) | 0.056 | -0.0171(-0.0324,-0.0018) | 0.028 |
| Subgroup analysis stratified by age |  |  |  |  |
| 18-45 | 0.0034(-0.0010,0.0078) | 0.126 | 0.0018(-0.0031,0.0066) | 0.478 |
| 45-65 | 0.0016(-0.0032,0.0064) | 0.517 | -0.0006(-0.0058,0.0047) | 0.832 |
| 65-80 | -0.0100(-0.0186,-0.0014) | 0.022 | -0.0144(-0.0234,-0.0054) | 0.002 |

Adjustments were made for age, sex, race/ethnicity (Mexican American, other Hispanic, non-Hispanic white, non-Hispanic black, other race), BMI, alcohol consumption, osteoporosis, COPD, thyroid problems, arthritis, and smoking. In subgroup analyses stratified by gender or race/ethnicity, the model was not adjusted for the stratification variables themselves.

**Table 4.** Association of blood chloroform with wards triangle BMD/BMC, stratified by sex, race/ethnicity and age.

| | Wards triangle BMD (g/cm<sup>2</sup>) | $\beta$(95%CI) value | Wards triangleBMC (g) | $\beta$(95%CI) value |
| --- | --- | --- | --- | --- |
| Subgroup analysis stratified by gender |  |  |  |  |
| male | -0.0020(-0.0110,0.0069) | 0.658 | -0.0023(-0.0118,0.0072) | 0.634 |
| female | -0.0118(-0.0204,-0.0032) | 0.007 | -0.0144(-0.0239,-0.0049) | 0.003 |
| Subgroup analysis stratified by race/ethnicity |  |  |  |  |
| Mexican American | -0.0024(-0.0148,0.0100) | 0.703 | -0.0036(-0.0172,0.0100) | 0.605 |
| Other Hispanic | -0.0068(-0.0244,0.0109) | 0.451 | -0.0090(-0.0281,0.0102) | 0.358 |
| Non-Hispanic White | -0.0124(-0.0224,-0.0024) | 0.015 | -0.0136(-0.0244,-0.0027) | 0.015 |
| Non-Hispanic Black | -0.0133(-0.0286,0.0021) | 0.091 | -0.0153(-0.0318,0.0012) | 0.070 |
| Other Race—Including Multi-Racial | -0.0103(-0.0293,0.0087) | 0.289 | -0.0118(-0.0324,0.0089) | 0.2639 |
| Subgroup analysis stratified by age |  |  |  |  |
| 18-45 | 0.0037(-0.0025,0.0099) | 0.241 | 0.0018(-0.0052,0.0088) | 0.616 |
| 45-65 | 0.0052(-0.0023,0.0128) | 0.175 | 0.0049(-0.0034,0.0131) | 0.251 |
| 65-80 | -0.0065(-0.0211,0.0081) | 0.381 | -0.0060(-0.0216,0.0095) | 0.447 |

Adjustments were made for age, sex, race/ethnicity (Mexican American, other Hispanic, non-Hispanic white, non-Hispanic black, other race), BMI, alcohol consumption, osteoporosis, COPD, thyroid problems, arthritis, and smoking. In subgroup analyses stratified by gender or race/ethnicity, the model was not adjusted for the stratification variables themselves.

**Table 5.** Association of blood chloroform with L4 BMD/BMC, stratified by sex, race/ethnicity and age.

| | L4BMD (g/cm<sup>2</sup>) | $\beta$(95%CI) value | L4BMC (g) | $\beta$(95%CI) value |
| --- | --- | --- | --- | --- |
| Subgroup analysis stratified by gender |  |  |  |  |
| male | -0.0012(-0.0057,0.0033) | 0.600 | -0.0034(-0.0098,0.0030) | 0.297 |
| female | -0.0044(-0.0090,0.0002) | 0.060 | -0.009(-0.0149,-0.0022) | 0.009 |
| Subgroup analysis stratified by race/ethnicity |  |  |  |  |
| Mexican American | -0.0055(-0.0125,0.0015) | 0.124 | -0.0107(-0.0217,0.0002) | 0.054 |
| Other Hispanic | -0.0028(-0.0124,0.0067) | 0.560 | -0.0058(-0.0204,0.0088) | 0.437 |
| Non-Hispanic White | -0.0044(-0.0093,0.0005) | 0.082 | -0.0078(-0.0152,-0.0005) | 0.037 |
| Non-Hispanic Black | -0.0066(-0.0142,0.0011) | 0.095 | -0.0109(-0.0223,0.0004) | 0.059 |
| Other Race—Including Multi-Racial | -0.0057(-0.0168,0.0054) | 0.313 | -0.0113(-0.0286,0.0059) | 0.196 |
| Subgroup analysis stratified by age |  |  |  |  |
| 18-45 | 0.0001(-0.0037,0.0039) | 0.957 | -0.0014(-0.0071,0.0043) | 0.625 |
| 45-65 | -0.0037(-0.0088,0.0014) | 0.156 | -0.0073(-0.0145,-0.0001) | 0.047 |
| 65-80 | -0.0072(-0.0162,0.0018) | 0.119 | -0.0154(-0.0282,-0.0026) | 0.018 |

Adjustments were made for age, sex, race/ethnicity (Mexican American, other Hispanic, non-Hispanic white, non-Hispanic black, other race), BMI, alcohol consumption, osteoporosis, COPD, thyroid problems, arthritis, and smoking. In subgroup analyses stratified by gender or race/ethnicity, the model was not adjusted for the stratification variables themselves.

Results of the multiple regression analysis of Ward’s triangle region revealed that stratified by gender, female Ward’s triangle BMD decreased while chloroform levels increased and that female Ward’s triangle BMC was negatively correlated with chloroform. According to the results stratified by race, blood chloroform concentration was negatively correlated with BMD/BMC except for Hispanic white subjects (β - 0.0124; 95%CI -0.0224, -0.0024; P<0.05); (β -0.0136; 95% CI -0.0244, -0.0027; P<0.05). Additionally, although BMC was not statistically significant, chloroform concentrations were correlated with BMD in Mexican-American, Other Hispanic, Non-Hispanic Black and Other Race-Including Multi-Racial participants. The results stratified by age revealed no statistically significant association between chloroform concentrations and BMD and BMC at all femoral sites in all age groups.

Female L4 BMC decreased with increasing blood chloroform in the spinal region following stratification by sex (β -0.009; 95% CI -0.0149, -0.0022; P<0.01). When stratified by race, we discovered a significant correlation between Non-Hispanic White BMC and blood chloroform (β -0.0078; 95% CI -0.0152, -0.0005; P<0.05). Additionally, there was a significant negative correlation between BMC and chloroform in the 45-65 and 65-80 age groups when age was used as a stratification factor (β -0.0073; 95% CI -0.0145, -0.0001; P<0.05); (β -0.0154; 95% CI -0.0282, - 0.0026; P< 0.05).

## Discussion

Osteoporosis is a degenerative bone disease that increases the risk of fracture [20]. With the aging population, osteoporosis is becoming a major public health concern worldwide [21,22]. The current era of rapid global economic development characterized by various industrial processes, including increased production of automobiles, tobacco, cleaning products, building materials, and other household products, has made VOCs more common in the human environment [23,24,25,26,27]. Several studies have demonstrated that some VOCs, such as Perfluoro and Polyfluoroalkyl Substances (PFAS), Urinary Polycyclic Aromatic Hydrocarbons (UPAHs), trichlorophenol, and many other organics are negatively associated with BMD/BMC levels at specific sites in the human body [28,29,30]. The safety of urban centralized water supply largely relies on water disinfection as it kills pathogens and prevents waterborne infectious diseases [31]. However, water Disinfection Byproducts (DBPs) mostly contain certain VOCs, including chloroform, bromine, dibromochloromethane, and dichlorobromomethane, with chloroform presence being exceptionally high in these DBPs [32]. A survey specifically focused on chloroform was conducted in China to evaluate the health risks associated with municipal water supply. It analyzed the distribution pattern and seasonal chloroform variation characteristics in the municipal water supply in Xi’an and the average daily chloroform exposure dose. According to the results, the average daily exposure dose of chloroform in the Xi’an municipal water supply was 7.39*10^-4 mg/(kg*d), with drinking water consumption as the leading source for the non-carcinogenic risk of chloroform [33]. Another survey found that the carcinogenic risk of ten contaminants in city drinking water, including chloroform, exceeded the acceptable level [34]. All these studies indicate that chloroform could potentially harm human health, but no conclusive study has proven the dangers of chloroform exposure to public safety. Although some studies have associated chloroform co-exposure with bone health indicators [15], no link with BMD has been reported. In our study, we hypothesize that there could be a connection between blood chloroform and BMD/BMC. After adjusting for confounders, we discovered a negative correlation between BMD/BMC and blood chloroform in L3 BMC and L4 BMD/BMC. Our analysis revealed a negative association between chloroform and femoral neck BMD in participants aged 65-80 years and Non-Hispanic White subjects, as well as Ward’s triangle BMD in women and Non-Hispanic White subjects. For BMC, a negative correlation was found between blood chloroform and femoral neck BMC, Non-Hispanic White and Other Race-Including Multi-Racial subjects femoral neck BMC, female Ward’s triangular BMC, Ward’s Triangular BMC of female Non-Hispanic White and 65-80 years old subjects, Female L4 and Non-Hispanic White Subjects BMC, and L4 BMC of 45-65 years and 65-80 years old subjects.

Multiple linear regression analysis and analysis within the Asian group revealed the relationship between blood chloroform and BMD/BMC in most areas of the femur and spine before adjusting for confounding factors. However, after adjusting for confounding variables, it was discovered that the correlation between blood chloroform and femoral BMD/BMC was not as strong. This finding implies that the relationship between blood chloroform and femoral BMD/BMC is largely influenced by age, gender, and race. On the one hand, this phenomenon may be attributed to femur and lumbar spine structures, which theoretically have the most substantial BMD changes in postmenopausal women with a higher proportion of cancellous bone [35, 36, 37, and 38]. On the other hand, it could be attributed to the sample population’s old age (48.8 ± 16.8 years), where hormonal drugs used by some older individuals exert a greater influence on femoral BMD/BMC than on lumbar spine BMD/BMC, making blood chloroform less likely to be associated with femoral BMD/BMC [39]. Furthermore, after adjusting for relevant covariates, stratified analysis by age, sex, and race revealed that chloroform concentration was negatively correlated with BMC at most sites. Blood chloroform concentration at L4 was significantly negatively correlated with BMC in women in the subgroup analysis. Contrastingly, no correlation between chloroform concentration and femoral or spinal BMC was observed in men, implying that blood chloroform levels may have some predictive value for bone mass in women. This phenomenon may be attributed to the sample population’s older age (48.8 ± 16.8 years) and the substantial decline in sex hormone levels in postmenopausal women resulting in lower femoral BMD quantity. Men, on the other hand, showed no such specific changes. We hypothesized that the varying sex hormone levels might influence the femoral neck, Ward’s triangle, or L4 BMC differently. Furthermore, the association between blood chloroform and femoral and spinal BMC was closer than that of BMD, which may be explained by the significant variations in the bone area resulting from individual differences.

Both BMD and BMC have been shown by studies to be significant osteoporosis indicators and to play a critical role in osteoporotic fracture development [40]. Our study revealed a negative correlation between blood chloroform in some spine regions and BMD/BMC. Chloroform is used as a solvent to soften material during endodontic treatment, and case studies revealed that during the patients’ endodontic retreatment visit, chloroform was extruded through the perforation into the periodontal ligament space, resulting in necrosis of the tooth-supporting bone [41]. Our findings on the negative effect of chloroform on bone loss are consistent with another research which discovered that organochlorine contaminants-exposure could impact fertility, the menstrual cycle, and other aspects of the human reproductive system [42,43]. In a pharmaceutical company, female personnel exposed to a mixture of organic solvents (including chloroform) exhibited higher Thyroid-Stimulating Hormone (TSH) levels than the non-exposed group, with a significant correlation between chloroform and TSH levels [44]. According to some clinical studies, a positive correlation exists between TSH and femoral and spinal BMD in early postmenopausal women [45]. Consequently, we speculate that chloroform may have a positive association with BMD/BMC mediated by hormones *in vivo*, and this hypothesis is supported by the “V”-shaped correlation between chloroform, and BMD/BMC discovered in our study (**Fig 2-3**). Additionally, we hope to find the link between chloroform and bone metabolism through preliminary data analysis and further basic research. In this regard, our group has already started collecting environmental exposure samples and establishing a sample bank. (**Fig2-3:**Restricted cubic spline plots between chloroform and femoral neck BMC and chloroform and L4 BMC)

With this cross-sectional research, we plan to further explore the effect of chloroform exposure on BMD/BMC. Herein, we used raw data on blood chloroform and femoral and spinal BMD/BMC from adults selected from NHANES in 2009-2010, 2013-2014, and 2017-2020 to perform a Restricted Cubic Splines (RCS) analysis of the relationship between chloroform and BMD/BMC. The non-linear connection between blood chloroform and spinal BMD/BMC revealed a “V” shaped correlation and a statistically significant association between chloroform levels within the 0-0.02 ng/ml range and spinal BMD/BMC (**Figure 2-3**). According to the literature review in some studies, chloroform harms BMD/BMC. Contrastingly, some studies suggest that chloroform may have a positive association with BMD/BMC. However, no specific reference in the literature demonstrates a clear connection between chloroform and BMD/BMC; thus, more research is required to determine the optimal level of chloroform exposure in relation to BMD/BMC levels. The above findings imply that exposure to VOCs such as chloroform remains a major public health concern. Disinfection byproducts are ubiquitous, and people (particularly older adults) with prolonged exposure to the chloroform from these products must be especially aware of their long-term risks.

### Limitation

Our study had several limitations. First, the data used in this investigation was obtained from NHANES, and therefore it was difficult to establish the exact timing between chloroform exposure and changes in BMD. Consequently, we could only develop correlations but not causality at this point. More research through prospective studies is therefore required to investigate this further. Second, our data on subjects were primarily from a single time point, making it difficult to determine the association between chloroform and BMD/BMC over time, potentially leading to bias in classifying specific populations. Third, some data were from self-reported questionnaires, which may have resulted in recall or misclassification bias. Fourth, missing whole-body skeletal BMD/BMC data impeded a comprehensive examination of the chloroform and BMD/BMC correlation. Finally, the residual confounding problem persisted despite employing multivariate adjustment techniques. We are developing a cohort of VOC exposure (including chloroform) in our city and expect that more convincing evidence elucidating the link between chloroform and BMD/BMC will soon be available.

## Conclusion

After adjustment for confounders, we established a “V” shaped correlation between blood chloroform and femoral spine BMD/ BMC, with a negative correlation at 0-0.02 ng/ml (p<0.05), especially for L4 BMC. Our research provides epidemiological evidence for further investigation into the correlation between chloroform and BMD/ BMC.

## Data Availability

All relevant data are within the manuscript and its Supporting Information files.

https://www.cdc.gov/nchs/nhanes/

## Acknowledgements

This thesis would not have been possible without the consistent and valuable reference materials that I received from my supervisor whose insightful guidance and enthusiastic encouragement in the course of my shaping this thesis definitely gains my deepest gratitude.

## Supporting information

**S1 Table Related demographic characteristics of participants included in NHANES from 2009 to 2010 and 2013 to 2014 and 2017-2020.** Means with standard deviation (SD) were used for continuous characteristic variables and categorical variables were expressed as frequencies. Differences in categorical variables between exposed groups were analyzed by Kruskal Wallis test or Mann Whitney test. Spearman correlation analysis was used to analyze the differences of continuous variables between groups. (EXCEL)

**S2 Table Blood chloroform and area, BMC, and BMD based on chloroform tristimulus.** Data are expressed as means (standard deviation, SD). BMC, bone mineral content, BMD, bone mineral density, Area (cm2), BMC (g), BMD (g/cm2). (EXCEL)

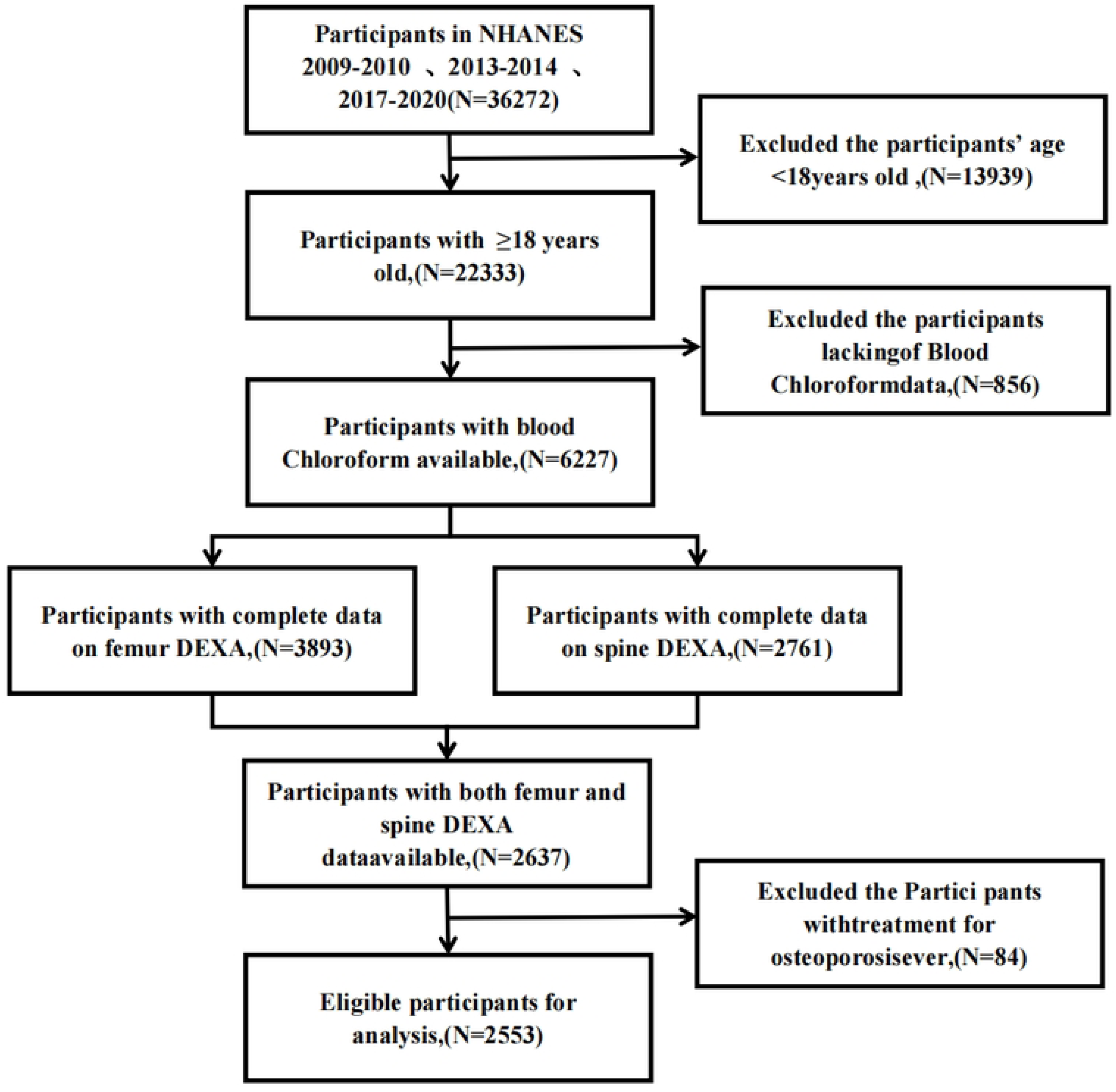

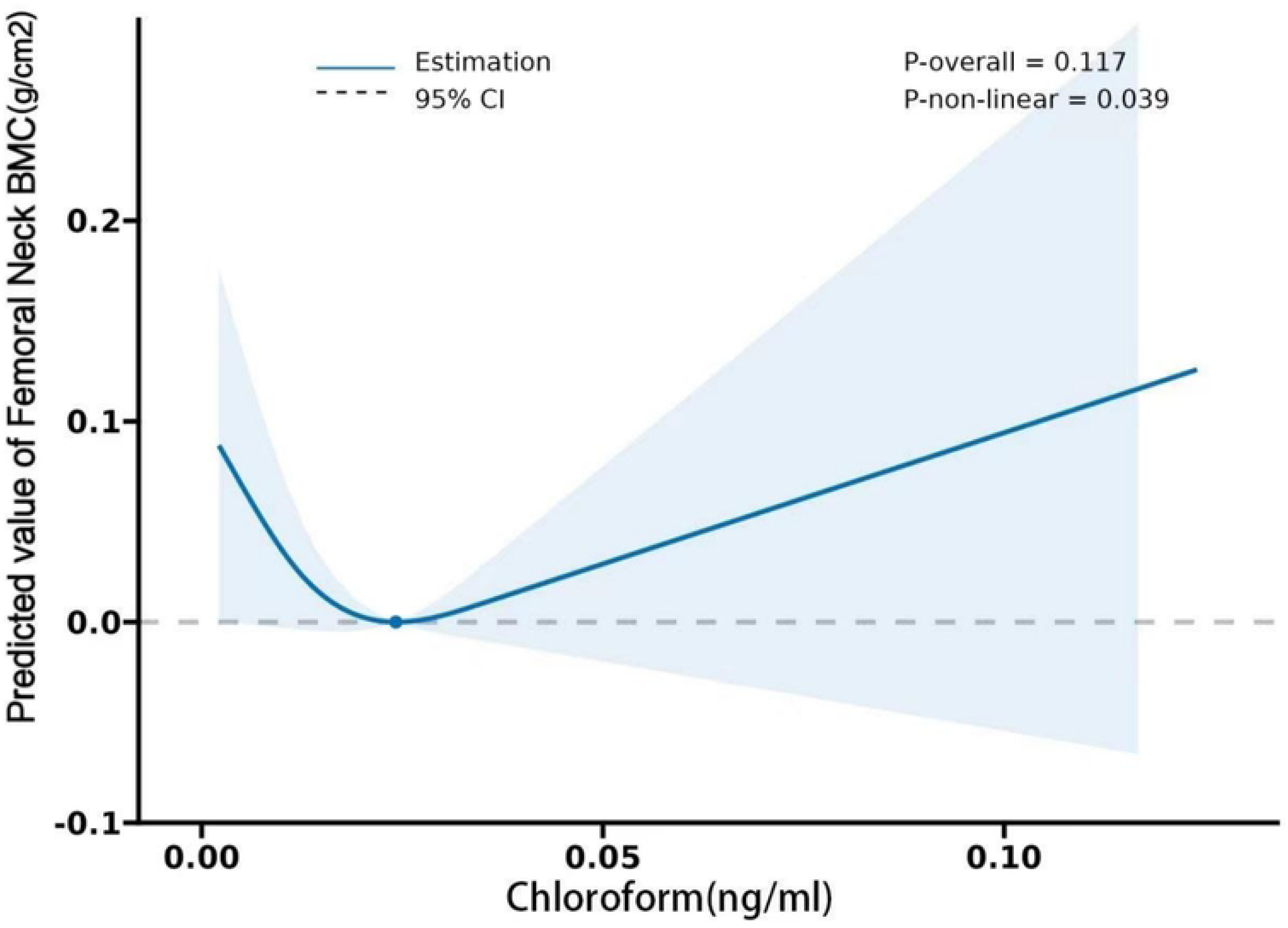

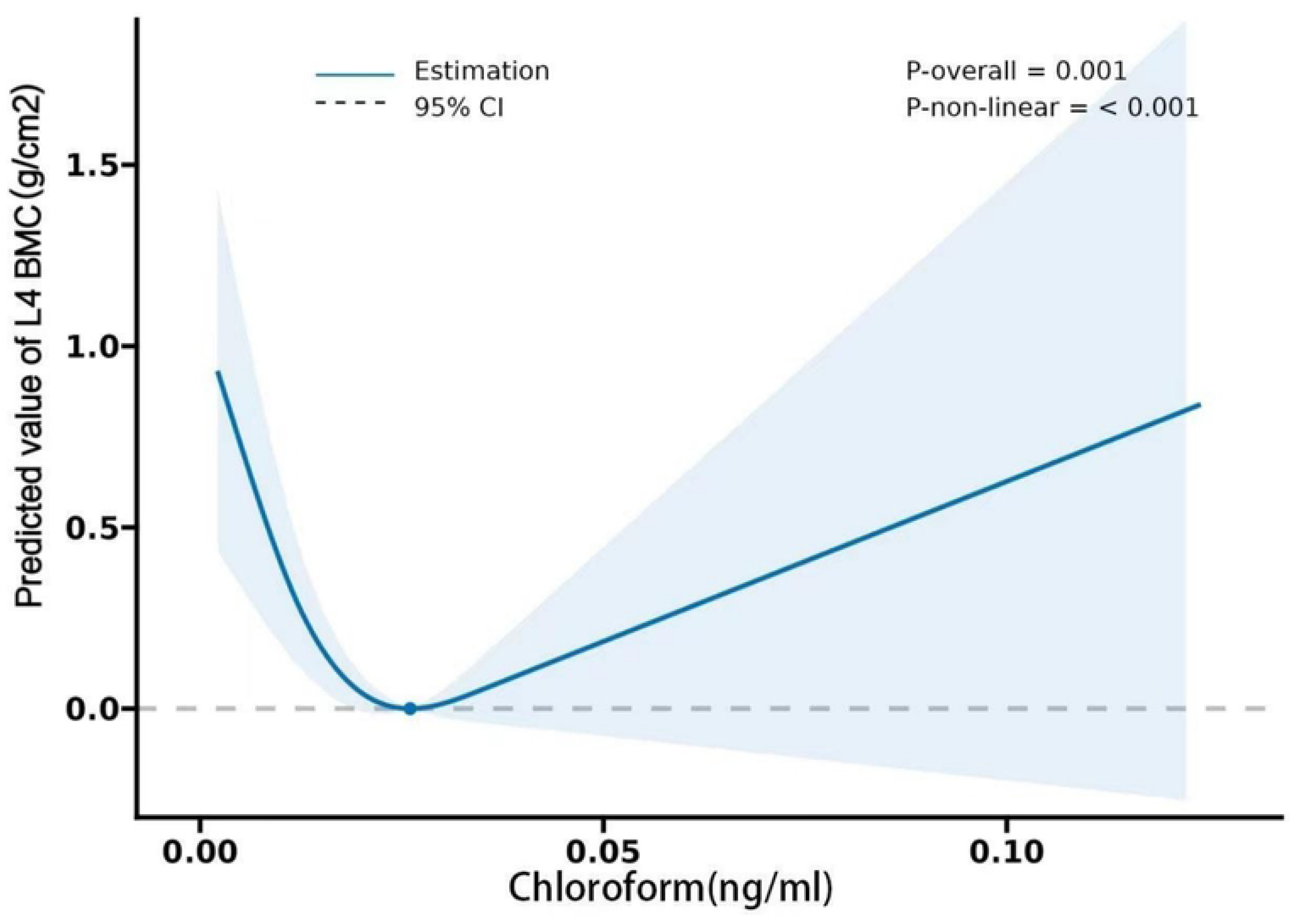

